# Who is lonely in lockdown? Cross-cohort analyses of predictors of loneliness before and during the COVID-19 pandemic

**DOI:** 10.1101/2020.05.14.20101360

**Authors:** Feifei Bu, Andrew Steptoe, Daisy Fancourt

**Author notes:** Corresponding author: Dr Daisy Fancourt, 1-19 Torrington Place, London, WC1E 7HB.

## Abstract

**Background:** There are concerns internationally that lockdown measures taken during the COVID-19 pandemic could lead to a rise in loneliness. As loneliness is recognised as a major public health concern, it is therefore vital that research considers the impact of the current COVID-19 pandemic on loneliness in order to provide necessary support. But it remains unclear who is lonely in lockdown?

**Methods:** This study compared socio-demographic predictors of loneliness before and during the COVID-19 pandemic using cross-cohort analyses of data from UK adults captured before the pandemic (UK Household Longitudinal Study, n=31,064) and during the pandemic (UCL COVID-19 Social Study, n=60,341).

**Results:** Risk factors for loneliness were near identical prior to and during the pandemic. Young adults, women, people with lower education or income, the economically inactive, people living alone, and urban residents had a higher risk of being lonely. Some people who were already at risk for being lonely (e.g. young adults aged 18-30, people with low household income, and adults living alone) experienced a heightened risk during the COVID-19 pandemic compared to prior to COVID-19 emerging. Further, being a student emerged as a higher risk factor during lockdown than usual.

**Conclusions:** Findings suggest that interventions to reduce or prevent loneliness during COVID-19 should be targeted at those socio-demographic groups already identified as high-risk in previous research. These groups are likely not just to experience loneliness during the pandemic but to have an even higher risk than normal of experiencing loneliness relative to low-risk groups.

## Introduction

Loneliness has is recognised as a major public health concern associated with heightened risk of mental and physical illness, cognitive decline, suicidal behaviour, and all-cause mortality ^1–3^. Loneliness itself has been referred to as an epidemic, and there have been heightened concerns about its effects during the global pandemic of COVID-19. Lockdowns and “stay-at-home” orders announced internationally have led to physical and social distancing and reports of many individuals experiencing social isolation. Whilst social isolation (the absence of social interactions, contacts and relationships with others) is conceptually distinguished from loneliness (the feeling that one’s social needs are not being met by the quantity or quality of one’s social relationships), the two are known to be inter-related, with isolation often being a risk factor for becoming lonely ^4^. As a result, there have been calls to ascertain how the pandemic has affected loneliness to ensure that individuals at risk receive necessary support ^5,6^.

In particular a key question is who is lonely in lockdown? On the one hand, individuals who already experience loneliness may be feeling even more isolated as a result of social distancing measures. Previous research has highlighted that particular groups at risk of loneliness include being female, being either younger (e.g. under 25) or older (e.g. over 65), living alone, low socio-economic status, and poor mental and physical health ^7,8^. Preliminary research within Europe has suggested that these groups may indeed be at risk during lockdown and heightened loneliness is also affecting distress levels ^9^. However, it is also possible that enforced lockdowns are actually meaning that new groups are now at risk of loneliness ^10^. The pandemic has forced millions globally to curtail face-to-face contact and social activities, cut jobs and employment opportunities, restrict travelling, and limit outdoor activity. For many individuals, this will be a radical departure from their patterns of usual daily life, and they may find habitual coping mechanisms (such as meeting with others) disrupted, leading to a heightened risk of feeling that the emotional and social support available to them is insufficient to meet their needs. Therefore, this study compared socio-demographic predictors of loneliness before and during the COVID-19 pandemic using cross-cohort analyses of data captured before and during the pandemic.

## Methods

### Participants

Data were drawn from two sources. For data collected prior to the pandemic, we used Understanding Society: the UK Household Longitudinal Study (UKHLS), a nationally representative household panel study of the UK population (2009-2019). Our analyses used the most recent wave of UKHLS (wave 9) where the loneliness measures were introduced. The wave 9 data were collected between January, 2017 and June, 2019. To be consistent with the UCL Covid-19 Social Study, we restricted participants to those aged 18+, leaving us a total sample size of 34,976 participants. Further, we excluded those who had missing value in loneliness or any of the covariates (11%). This provided a final sample size of 31,064.

For data during the COVID-19 pandemic, we used data from the UCL COVID-19 Social Study; a large panel study of the psychological and social experiences of over 50,000 adults (aged 18+) in the UK. The study commenced on 21^st^ March 2020 involving online weekly data collection from participants for the duration of the COVID-19 pandemic in the UK. Whilst not random, the study has a well-stratified sample that was recruited using three primary approaches. First, snowballing was used, including promoting the study through existing networks and mailing lists (including large databases of adults who had previously consented to be involved in health research across the UK), print and digital media coverage, and social media. Second, more targeted recruitment was undertaken focusing on (i) individuals from a low-income background, (ii) individuals with no or few educational qualifications, and (iii) individuals who were unemployed. Third, the study was promoted via partnerships with third sector organisations to vulnerable groups, including adults with pre-existing mental illness, older adults, and carers. The study was approved by the UCL Research Ethics Committee [12467/005] and all participants gave informed consent. In this study, we focused on participants who had a baseline response between 21^st^ March and 10^th^ May 2020. This provided us with data from 67,142 participants. Of these, 10% participants withheld data on socio-demographic factors including gender and income so were excluded, providing a final analytic sample size of 60,341.

### Measures

In both datasets, loneliness was measured using the three-item UCLA loneliness scale (UCLA-3). The questions include: 1) how often do you feel lack companionship? 2) how often do you feel isolated from others? 3) how often do you feel left out? Responses to each question were scored on a three-point Likert scale ranging from hardly ever/never, to some of the time, to often. Using the sum score, we had a loneliness scale ranging from 3 to 9, with a higher score indicating increased loneliness. In addition, we also examined the single-item direct measure of loneliness asking how often the respondent felt lonely, which was coded on the same scale as the UCLA-3 items.

Covariates included age groups (18-29, 30-45, 46-59 and 60+), gender (woman vs. man), ethnicity (non-white vs. white), education (low: GCSE or below, medium: A levels or equivalent, high: degree or above), low income (household annual income <£30,000 vs higher household annual income), employment status (employed, unemployed, student and inactive other), living status (alone, with others but no children, with others including children) and area of living (rural vs. urban). All variables above were harmonised between the two datasets.

### Analysis

To compare risk factors for loneliness, we used Ordinary Least Square (OLS) regression models fitted separately in the two datasets. Survey weights were applied to both samples throughout the analyses to yield national representative samples of UK adults. The analyses of UKHLS used cross-sectional adult self-completion interview weights while analyses of the UCL Covid-19 Social study were weighted to the proportions of gender, age, ethnicity, education and country of living obtained from the Office for National Statistics ^11^. The descriptive and regression analyses were implemented in Stata v15 (StataCorps, Texas).

## Results

Descriptive statistics for the two samples are shown in Table 1. Loneliness levels were higher in the UCL COVID-19 Social Study than in UKHLS, with 32.5% of people feeling lonely sometimes (28.6% in UKHLS) and 18.3% often (8.5% in UKHLS).

Risk factors for loneliness were near identical prior to and during the pandemic (Figure 1). Adults aged 18-30 were more likely to be lonely compared to adults aged 60+ prior to the pandemic (coef=1.01, 95%CI 0.89-1.12), and during the pandemic (coef=1.58, 95%CI 1.48-1.68). People living alone, similarly, were more at risk prior to and during the pandemic ((coef=0.61, 95%CI 0.51-0.71 vs coef=1.10, 95%CI 1.02-1.18). Having a low household income and being unemployed were also persistent risk factors. Being a student was only a moderate risk factor prior to the pandemic (coef=0.19, 95%CI 0.02-0.35), but was a greater risk factor during the pandemic (coef=0.43, 95%CI 0.28-0.58). Other risk factors including non-white ethnicity, being female, low educational attainment, and living in urban areas were only small risk factors but were maintained before and during the pandemic.

**Table 1.**
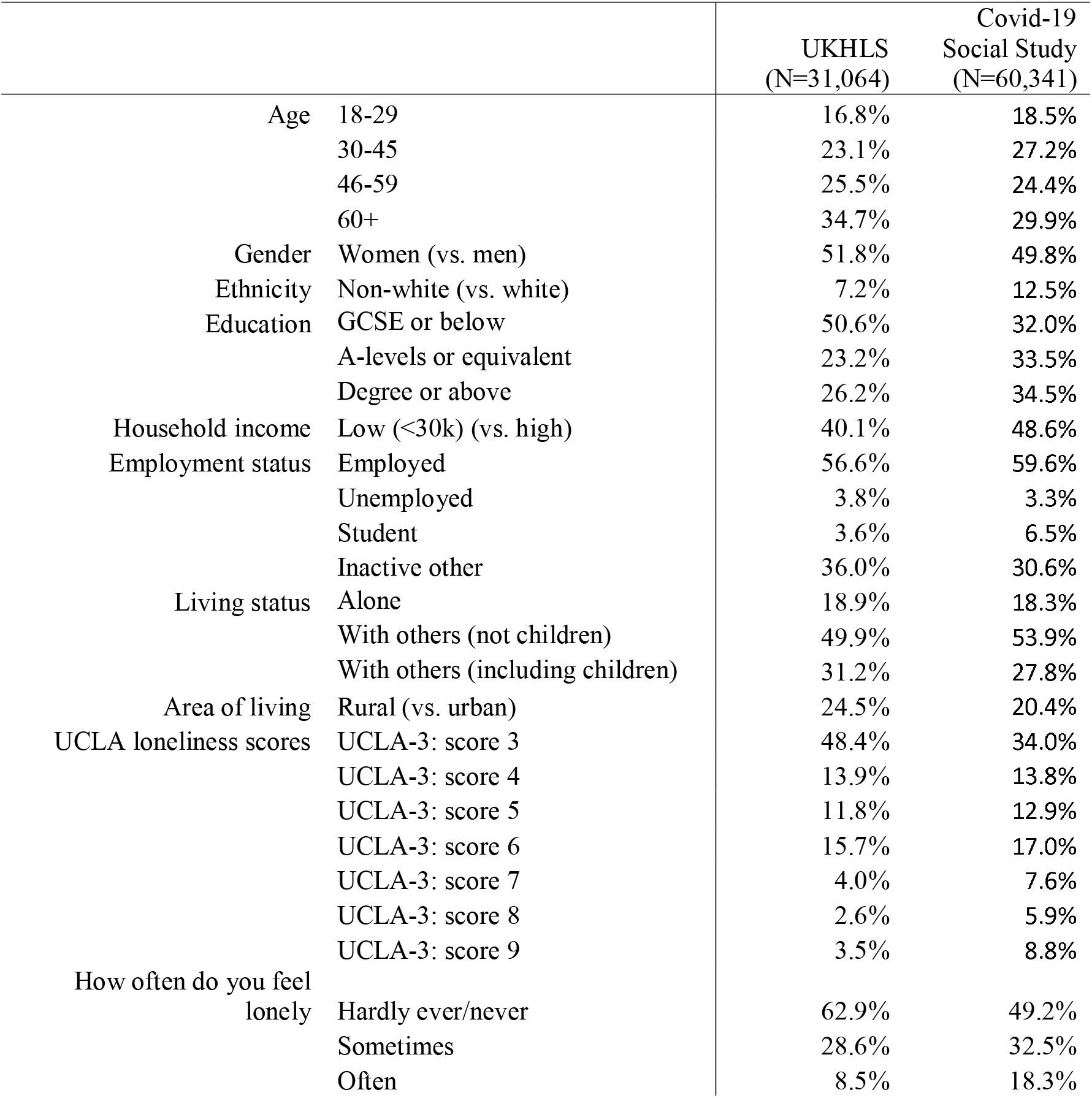
Descriptive statistic of the explanatory variables (weighted)

**Figure 1.**
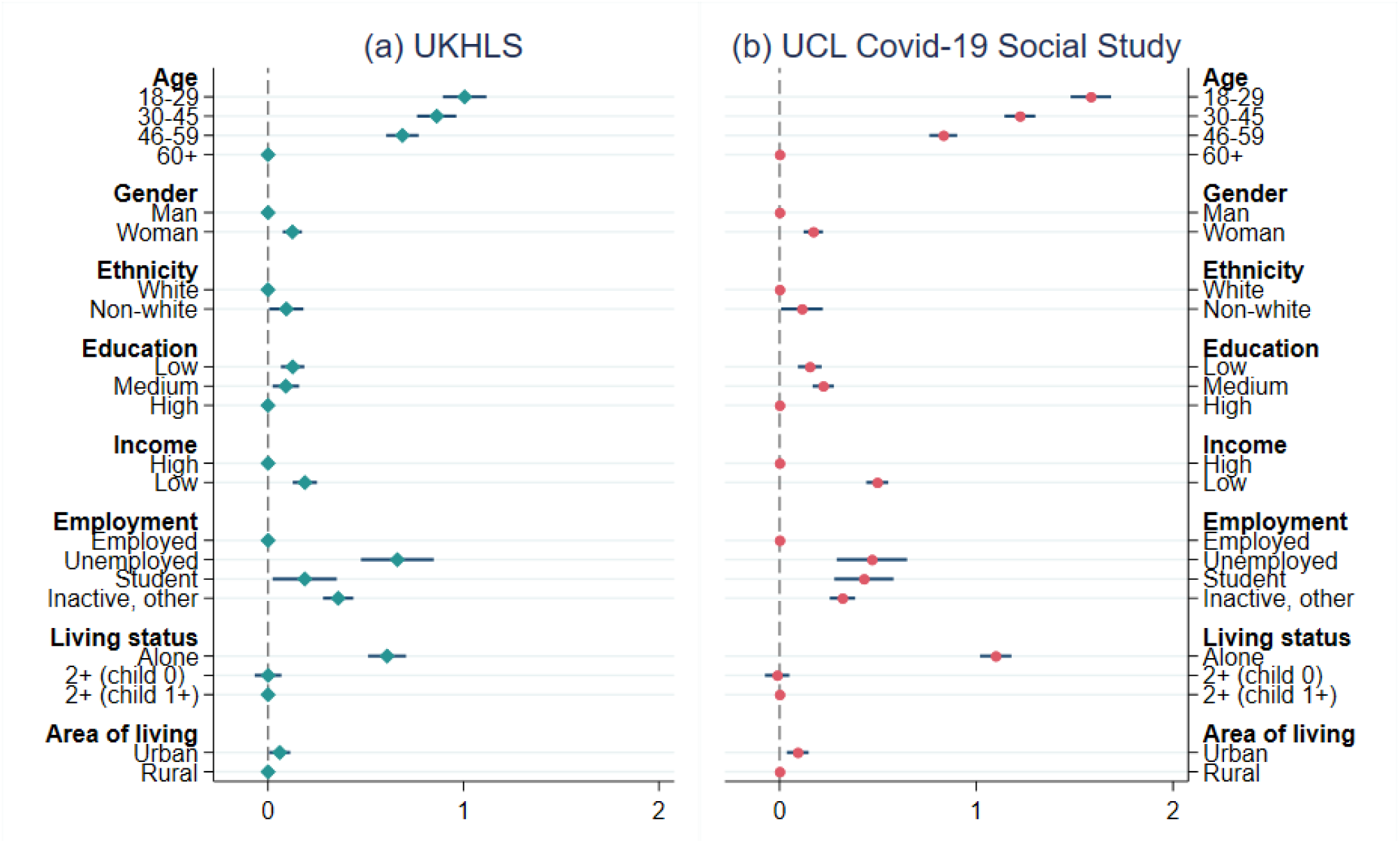
Coefficients and 95% confidence intervals from the regression model on loneliness

## Discussion

This study explored who was most at risk of loneliness during the UK lockdown due to the COVID-19 pandemic and compared whether risk factors were similar to risk factors for loneliness before the pandemic. Young adults, people living alone, people with lower education or income, the economically inactive, women, ethnic minority groups, and urban residents had a higher risk of being lonely both prior to and during the pandemic. These results echo previous studies on risk factors for loneliness ^7,8^. These findings in the UK are also echoed by some recent data from Spain during their lockdown, which highlighted similar risk factors ^9^. However, these data show that some people who were already at risk for being lonely (e.g. young adults aged 18-30, people with low household income, and adults living alone) experienced an even greater risk during the COVID-19 pandemic compared to usual (indicated by higher coefficients). Further, being a student emerged as a higher risk factor during lockdown than usual, although this builds on wider research suggesting that loneliness can be a problem for students and has been rising over the past 6 years ^12^.

This study has a number of strengths including its cross-cohort comparisons of two large samples with harmonised measures before and during the pandemic, and its consideration of a broad range of socio-demographic characteristics. However, the data compared are from different participants, so it is not clear whether those individuals experiencing loneliness during lockdown had previous experience of loneliness. Further, the COVID-19 Social Study is a non-random (albeit large, heterogeneous, well-stratified, and weighted) sample. So the results presented here are not presented as accurate prevalence figures for loneliness during the pandemic. It is possible that the study inadvertently attracted individuals who were feeling more lonely to participate. Finally, the study looked at broad risk categories. Future studies are encouraged that (i) consider whether the interaction between different risk categories (e.g. unemployed adults living alone) or accumulation of multiple risk factors affected loneliness levels during the pandemic, (ii) track the trajectories of loneliness across lockdown, and (iii) explore the potential buffering role of protective social or behavioural factors.

Overall, these findings suggest that interventions to reduce or prevent loneliness during COVID-19 should be targeted at those socio-demographic groups already identified as high-risk in previous research. These groups are likely not just to experience loneliness during the pandemic but to have an even higher risk than normal of experiencing loneliness relative to low-risk groups. Such efforts are particularly important given rising concerns that loneliness could exacerbate mental illness and lead to non-adherence to government regulations ^13,14^. As such, supporting individuals experiencing loneliness during and in the aftermath of the pandemic should be a public health priority.

## Data Availability

Anonymous data will be made available following the end of the pandemic.

## References

1. Jeste DV, Lee EE, Cacioppo S. Battling the Modern Behavioral Epidemic of Loneliness: Suggestions for Research and Interventions. JAMA Psychiatry. Published online March 4, 2020. doi:10.1001/jamapsychiatry.2020.0027

2. Leigh-Hunt N, Bagguley D, Bash K, et al. An overview of systematic reviews on the public health consequences of social isolation and loneliness. Public Health. 2017;152:157–171. doi:10.1016/j.puhe.2017.07.035

3. Stickley A, Koyanagi A. Loneliness, common mental disorders and suicidal behavior: Findings from a general population survey. Journal of Affective Disorders. 2016;197:81–87. doi:10.1016/j.jad.2016.02.054

4. Hawkley LC, Cacioppo JT. Loneliness Matters: A Theoretical and Empirical Review of Consequences and Mechanisms. Ann Behav Med. 2010;40(2):218–227. doi:10.1007/s12160-010-9210-8

5. Armitage R, Nellums LB. COVID-19 and the consequences of isolating the elderly. The Lancet Public Health. 2020;5(5):e256. doi:10.1016/S2468-2667(20)30061-X

6. Banerjee D, Rai M. Social isolation in Covid-19: The impact of loneliness. Int J Soc Psychiatry. Published online April 29, 2020:0020764020922269. doi:10.1177/0020764020922269

7. Pinquart M, Sörensen S. Risk factors for loneliness in adulthood and old age--a meta-analysis. In: Advances in Psychology Research, Vol. 19. Nova Science Publishers; 2003:111–143.

8. Victor CR, Yang K. The Prevalence of Loneliness Among Adults: A Case Study of the United Kingdom. The Journal of Psychology. 2012;146(1-2):85–104. doi:10.1080/00223980.2011.613875

9. Losada-Baltar A, Jiménez-Gonzalo L, Gallego-Alberto L, Pedroso-Chaparro M del S, Fernandes-Pires J, Márquez-González M. “We Are Staying at Home.” Association of Self-perceptions of Aging, Personal and Family Resources, and Loneliness With Psychological Distress During the Lock-Down Period of COVID-19. J Gerontol B Psychol Sci Soc Sci. doi:10.1093/geronb/gbaa048

10. Fancourt D, Steptoe A. Is this social isolation?—we need to think broadly about the impact of social experiences during covid-19. The BMJ. Published May 22, 2020. Accessed June 12, 2020. https://blogs.bmj.com/bmj/2020/05/22/is-this-social-isolation-we-need-to-think-broadly-about-the-impact-of-social-experiences-during-covid-19/

11. Overview of the UK Population: November 2018. Office for National Statistics; 2018. Accessed May 7, 2020. https://www.ons.gov.uk/releases/overviewoftheukpopulationnovember2018

12. Hysing M, Petrie KJ, Bøe T, Lønning KJ, Sivertsen B. Only the Lonely: A Study of Loneliness Among University Students in Norway. 1. 2020;2(1):1–16. doi:10.32872/cpe.v2i1.2781

13. Okruszek L, Aniszewska-Stańczuk A, Piejka A, Wiśniewska M, Żurek K. Safe but Lonely? Loneliness, Mental Health Symptoms and COVID-19. PsyArXiv; 2020. doi:10.31234/osf.io/9njps

14. Cerami C, Santi GC, Galandra C, et al. COVID-19 Outbreak in Italy: Are We Ready for the Psychosocial and Economic Crisis? Baseline Findings from the Longitudinal PsyCovid Study. Social Science Research Network; 2020. Accessed May 5, 2020. https://papers.ssrn.com/abstract=3569868

